# Psychometric profiling of patients with chronic gastroduodenal symptoms using body surface gastric mapping phenotypes

**DOI:** 10.1101/2025.07.28.25332341

**Authors:** Mikaela Law, Gabriel Schamberg, Chris Varghese, Billy Wu, Charlotte Daker, Isabella Pickering, Gen Johnston, Daphne Foong, Vincent Ho, Christopher N. Andrews, Armen Gharibans, Greg O’Grady, Stefan Calder

**Author notes:** Correspondence: Dr Stefan Calder, Department of Surgery, The University of Auckland, Auckland, New Zealand. Joint Senior Authors (Greg O’Grady, Stefan Calder).

## Abstract

**Background and aims:** Chronic gastroduodenal symptoms may be associated with psychological factors; however, recent evidence suggests these associations vary by previously undetermined disease factors. Body surface gastric mapping (BSGM) is a non-invasive diagnostic method integrating high-resolution myoelectrical measurement and validated symptom profiling. This cross-sectional study investigated associations between psychological factors and BSGM phenotypes.

**Methods:** Patients from the general community meeting the Rome IV Criteria for functional dyspepsia or chronic nausea and vomiting syndrome underwent BSGM using Gastric Alimetry^®^. The test protocol included a 30-min fasting baseline, 482 kCal meal, and 4-hr postprandial recording. Measures of depression, anxiety, stress, and quality of life were assessed at baseline, and symptoms were logged throughout the test. BSGM phenotypes were classified using established rule-based criteria.

**Results:** Among 278 patients (mean age 39.5, 15-88; 77% female), clinical diagnoses of depression (45%) and anxiety (46%) were common. Depression, anxiety, and stress measures were positively associated with symptom severity; however, these associations varied substantially by BSGM phenotype. Abnormal rhythm stability predicted higher depression (*B*=0.35, *p*=.044) and stress (*B*=1.48, *p*=.026). Among patients with normal spectral metrics, continuous symptoms predicted higher levels of depression (*B*=0.42, *p*=.003), anxiety (*B*=0.30, *p*=.045), and stress (*B*=1.43, *p*=.008), and worse quality of life (*B*=-0.57, *p*< .001); while sensorimotor symptoms predicted higher anxiety (*B*=0.46, *p*=.029) and worse quality of life (*B*=-0.49, *p*=.033).

**Conclusion:** This study confirms significant connections between gastroduodenal symptoms and mental health, but refines these associations to specific BSGM phenotypes. Individuals exhibiting normal spectral metrics alongside continuous or sensorimotor symptoms may particularly benefit from integrated psychological interventions.

## Introduction

Chronic gastroduodenal symptoms, which are hallmarks of functional dyspepsia (FD) and chronic nausea and vomiting syndrome (CNVS), place a substantial strain on patients and healthcare systems globally^1,2^. Due to heterogeneous pathophysiologies and overlapping symptomatology, affected patients often endure extensive testing and trial-and-error treatment, often with little effective symptom relief despite high healthcare utilization^3,4^. Improved diagnostic pathways and targeted therapies are needed to advance patient care.

Gastroduodenal disorders are multifactorial, with biopsychosocial models emphasizing the complex interplay of biological, social, and psychological factors in the onset, exacerbation, and maintenance of symptoms^5,6^. Psychological comorbidities are prevalent, with depression, anxiety, and stress being highly correlated with symptom severity, likely mediated via the gut-brain axis^7,8^. Consequently, psychological interventions, including cognitive behavioral therapy (CBT) and hypnotherapy, have shown promise in improving both mental health and gastrointestinal symptoms in clinical studies^9–11^. However, current frameworks, including the Disorders of Gut–Brain Interaction (DGBI) nomenclature, may inadvertently imply a uniform role for psychological factors across all patients, potentially obscuring important physiological heterogeneity^12^.

Recent translational advances are enabling a more granular understanding of the mechanisms of gastroduodenal symptoms^13,14^. One method to categorize these patients is body surface gastric mapping (BSGM), a new non-invasive test of gastric function that integrates high-resolution myoelectrical measurement with concurrent, validated symptom profiling^14–16^. This technology enables detailed characterization of patients with FD or CNVS into disease mechanism-specific subgroups (‘phenotypes’)^14,16–19^. Recent studies have indicated that BSGM phenotypes may reflect meaningful physiological differences that can inform diagnosis, reduce reliance on more generic labels, and improve healthcare utilization^14,20–22^.

Preliminary evidence using BSGM has revealed links between psychological factors and specific BSGM test phenotypes^14,17,19,23,24^. Specifically, patients with normal gastric myoelectrical activity have been found to report higher depression and anxiety levels compared to those with detectable gastric dysfunction in preliminary studies^14^. These data have helped to inform an emerging BSGM classification scheme that could improve diagnostic specificity and targeted care^25–27^. This study, therefore, now aimed to comprehensively investigate the association between psychological factors and the spectral metrics, symptom patterns, and phenotypes derived from BSGM in patients with FD and CNVS in a large cohort.

## Materials and Methods

Data were collected in Auckland (New Zealand), Calgary (Canada), and Western Sydney (Australia) as part of an international BSGM consortium study. Ethical approvals were granted by the Auckland Health Research Ethics Committee (AHREC; AH1130 & AH27068), the University of Calgary Conjoint Health Research Ethics Board (REB19-1925), and the Human Research Ethics Committee at Western Sydney (H15157). All participants provided written informed consent.

Patients with chronic gastroduodenal symptoms were recruited from the general community to complete a standard BSGM test using Gastric Alimetry® (Alimetry, New Zealand) alongside a battery of psychometrics as part of a multi-national consortium database study, on behalf of the BSGM Consortium. Patients were included if they met the Rome IV symptom Criteria for FD or CNVS^28^. Exclusion criteria included pregnancy or breastfeeding, previous major gastric surgery, as well as standard cautions against Gastric Alimetry testing (e.g. adhesive allergies or damaged epigastric skin)^25^. All tests occurred between June 2022 and September 2024.

### Procedure

Participants underwent a Gastric Alimetry test following standardized procedures^14,16,18,25^. Tests were conducted in the morning after an overnight fast of at least six hours. Participants abstained from taking medications affecting gastrointestinal motility for 48 hours before the test, including promotility agents and opiates, and avoided caffeine and nicotine on the test day^18^. The Gastric Alimetry system consists of a stretchable 8×8 electrode array on an adhesive patch and a wearable reader, which is placed over the epigastrium. After a 30-minute fasting baseline recording, participants consumed a standardized 482 kcal test meal, followed by a 4-hour postprandial recording^18^. The standard meal was an oatmeal energy bar (250 kcal, 5 g fat, 45 g carbohydrate, 10 g protein, 7 g fiber; Clif Bar & Company, CA, USA) and Ensure nutrient drink (232 kcal, 250 mL; Abbott Nutrition, IL, USA), or a calorie-matched diabetic or gluten-free alternative.

### Measures

#### BSGM metrics

A spectrogram is generated for each test, presenting the frequency and amplitude of gastric activity over time^16^. Three spectral metrics are then calculated^25,29,30^:

- *BMI-Adjusted Amplitude (µV):* characterizes the strength of the recorded gastric activity as an average of the whole-test amplitude, adjusted for body mass index (BMI)
- *Gastric Alimetry Rhythm Index™ (GA-RI):* a measure of the concentration of amplitude within the gastric frequency band over time (between 0-1), reflecting the rhythmic stability of gastric activity (also adjusted for BMI)
- *Principal Gastric Frequency (cpm):* characterizes the frequency associated with stable, consistent gastric activity (as defined by the GA-RI)

#### Gastroduodenal symptoms

Participants used the validated Gastric Alimetry App to record their gastroduodenal symptoms during the test^15^. Nausea, bloating, upper gut pain, heartburn, stomach burn, and excessive fullness were rated every 15 minutes on a scale of 0 (none) to 10 (most severe imaginable). Early satiety was assessed once after the meal, on the same scale. Average symptom scores during the test were calculated for each continuous symptom. A total symptom burden score was calculated by summing the average symptom scores, plus the early satiety score^15^. The app also recorded symptom events for reflux, belching, and vomiting.

#### Psychometrics

The following psychometrics were measured during the fasted baseline period: the Patient Health Questionnaire 9 (PHQ-9)^31^ to measure depression, the Generalized Anxiety Disorder 7 (GAD-7)^32^ to measure anxiety, the Perceived Stress Scale 4 (PSS-4)^33^ to measure chronic stress, and the Patient Assessment of Upper GastroIntestinal Disorders-Quality of Life

(PAGI-QoL) to measure quality of life^34^. Patients also self-reported whether they had a clinical diagnosis of anxiety or depression.

### Data analysis

All patients were grouped based on their BSGM metrics and symptom profiles using rule-based criteria, aligning with emerging phenotypes currently undergoing further refinement (see Figure 1)^26,27^. Firstly, patients were independently categorized into distinct spectral phenotypes based on the normative reference intervals of the three BSGM metrics^25,30^. Gastric dysfunction was categorized as low rhythm stability (GA-RI <0.25), low amplitude (BMI-Adjusted Amplitude <22 μV), high amplitude (>70 μV), high frequency (>3.35 cpm), and low frequency (<2.65 cpm)^25^. If a patient did not fall outside the normative reference intervals for all three metrics, they were classified as ‘normal on the day’.

**Figure 1.**
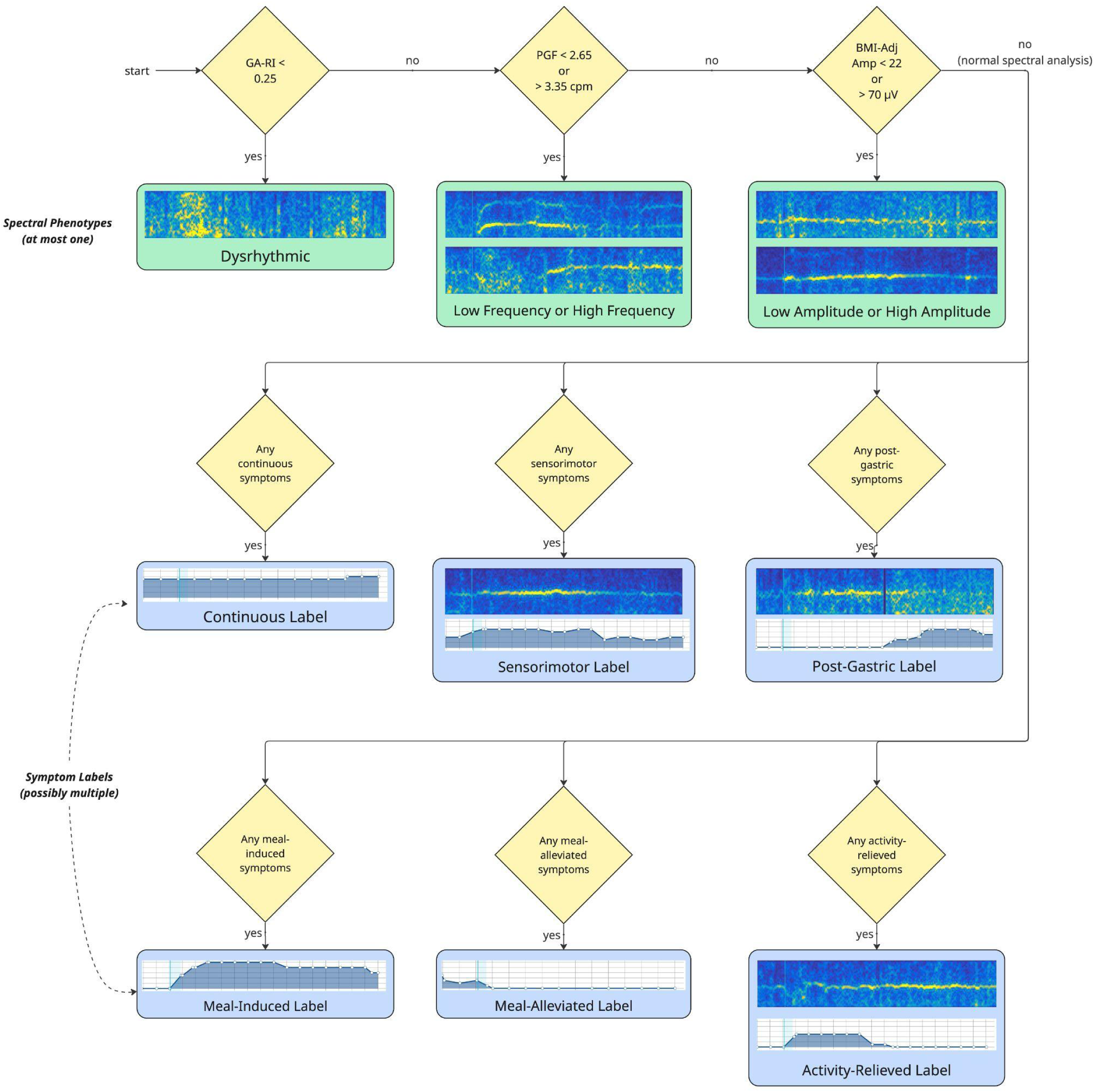
Flow diagram representing the BSGM phenotypes. First, tests are assessed for the presence of a spectral phenotype (green boxes). Tests with normal spectral analyses proceed to symptom-based classifications, with potentially multiple symptom labels (blue boxes) assigned to a single test. Spectrograms and symptom curves provide a representative example from the cohort.

Subsequently, patients with normal spectral metrics were further classified using symptom profiles if any of their symptoms measured from the validated Gastric Alimetry app met any of the following symptom patterns, described in detail elsewhere^27,30^. Six symptom profiles were applied here to comprehensively explore potential clinical relationships^35–37^.

- *Continuous:* symptom remains elevated (> 2/10) with minimal variation (range< 3) so as not to show variation induced by a meal stimulus or gastric motility
- *Sensorimotor:* strong correlation (r> .5) between the symptom severity and gastric amplitude
- *Post-gastric*: symptom severity increases after postprandial gastric activity has concluded
- *Meal-induced:* symptom severity increases after meal consumption: average 1st hr post-meal symptoms - pre-meal symptoms > 2
- Meal-alleviated: symptom severity decreases after meal consumption: average 1st hr post-meal symptoms - pre-meal symptoms < -2
- *Activity-relieved:* symptom severity decreases with increased gastric response

Patients could be assigned multiple symptom profiles if they had symptoms with differing patterns.

### Statistical analysis

Data were analyzed using IBM SPSS Statistics v29. Statistical significance was set to *p*<.05. Means (*M*) and standard deviations (*SD*) were reported unless otherwise stated. The PHQ-9 and GAD-7 total scores were transformed using a natural log transformation to address normality violations and logged values were used in all subsequent analyses.

Pearson’s correlations were conducted to assess the relationships between psychometric scores, symptoms, and BSGM metrics, both for the overall sample and within distinct Rome diagnostic groups (FD, CNVS, postprandial distress syndrome (PDS), and epigastric pain syndrome (EPS) alone, and overlapping CNVS/FD). Independent samples t-tests were performed to quantify the differences in the symptoms and spectral metrics between those with and without a clinical diagnosis of anxiety and depression. One-way ANOVAs were used to evaluate psychometric differences among the Rome diagnostic groups.

Independent multivariable linear regressions were conducted to assess the associations between the spectral phenotypes and symptom profiles, and the psychometrics. The spectral phenotypes were dummy-coded in reference to the ‘normal on the day’ phenotype. Symptom profiles were coded as 1 in those with normal spectral metrics if any symptom experienced during the test met the relevant symptom pattern. These associations were adjusted for age, sex, and BMI, and outputs were reported with 95% confidence intervals to assess statistical significance. Four independent regression models were performed, one for each outcome variable (lnPHQ-9 total score, lnGAD-7 total score, PSS-4 total score, and PAGI-QoL total score).

## Results

### Sample characteristics

The sample comprised 278 patients (*M* age 39.52 years, range 15-88 years; 77% female; *M* BMI 26.14). The majority of the sample (n=193, 69%) met Rome criteria for both CNVS and FD. Only a smaller proportion met criteria for FD alone (n=73, 26%) or CNVS alone (n=12, 4%). Gastric emptying data were not collected. On average, the sample reported moderate levels of depression (*M*= 11.02, *SD*= 6.80), mild levels of anxiety (*M*= 6.72, *SD*= 5.56), and moderate levels of stress (*M*= 7.29, *SD*= 3.31). Additionally, nearly half self-reported a clinical diagnosis of depression (45%) and anxiety (46%).

### Symptom-psychometric relationships

As shown in Figure 2, Pearson’s correlations revealed significant positive associations between most symptoms and psychometric scores, with the strongest effect sizes observed for quality of life and depression. Total symptom burden, nausea, upper gut pain, and stomach burn showed the strongest associations.

**Figure 2.**
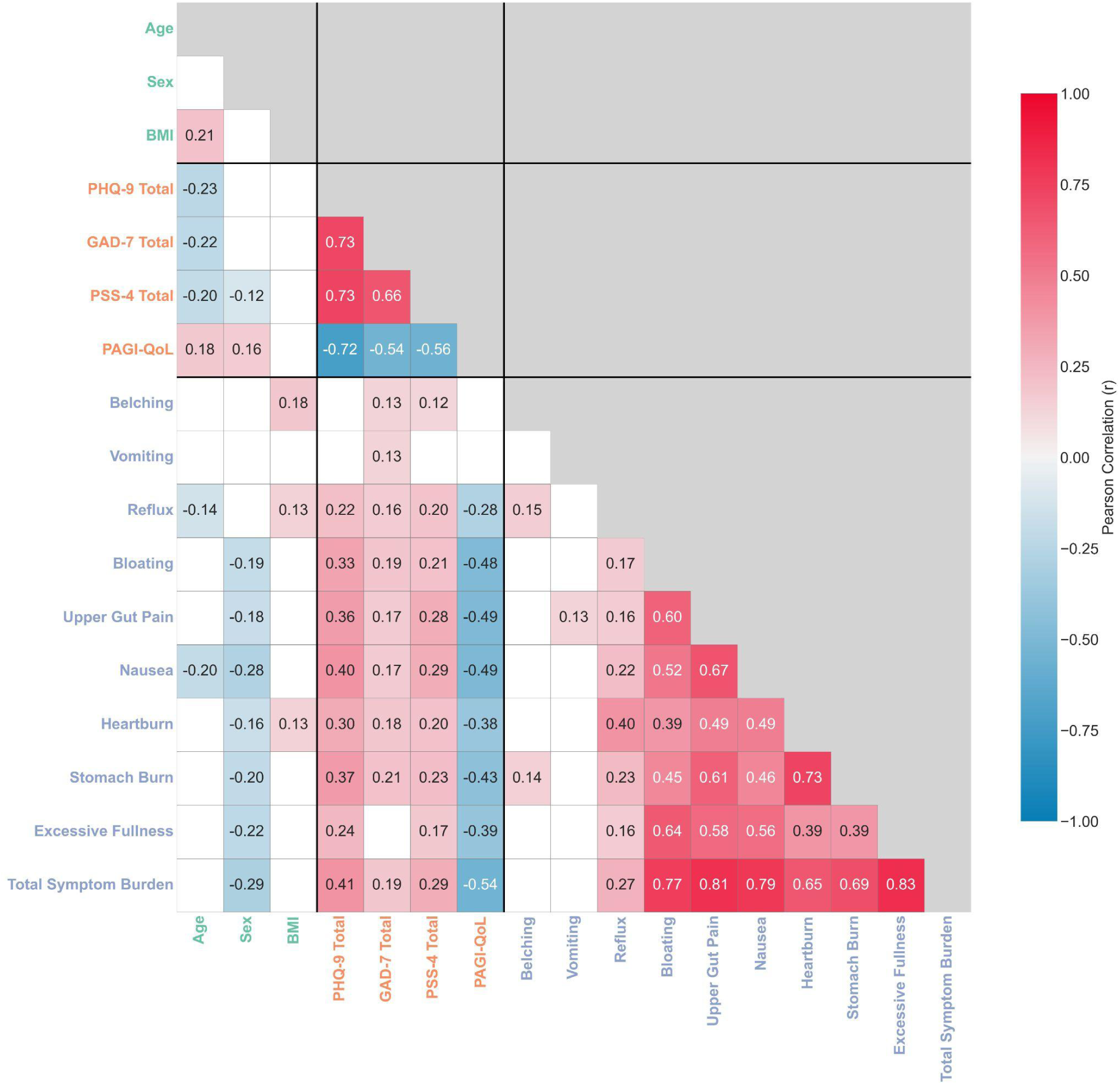
Correlation matrix showing the significant Pearson correlation coefficients between the psychometric scales (orange), symptom scores (purple), and demographic variables (green). Significance was determined using a Benjamini-Hochberg correction for multiple comparisons with a false discovery rate of 0.1.

Furthermore, as shown in Table 1, independent-sample t-tests revealed that patients with a self-reported clinical diagnosis of anxiety or depression experienced significantly higher symptom levels compared to those without such a diagnosis. The strongest effects were observed in patients with a depression diagnosis. Specifically, nausea showed the largest effect sizes for both depression (*d*= −0.47) and anxiety (*d*= −0.42). Upper gut pain, bloating, heartburn, reflux, and total symptom burden also demonstrated moderate effect sizes (all *d*>

**Table 1.**
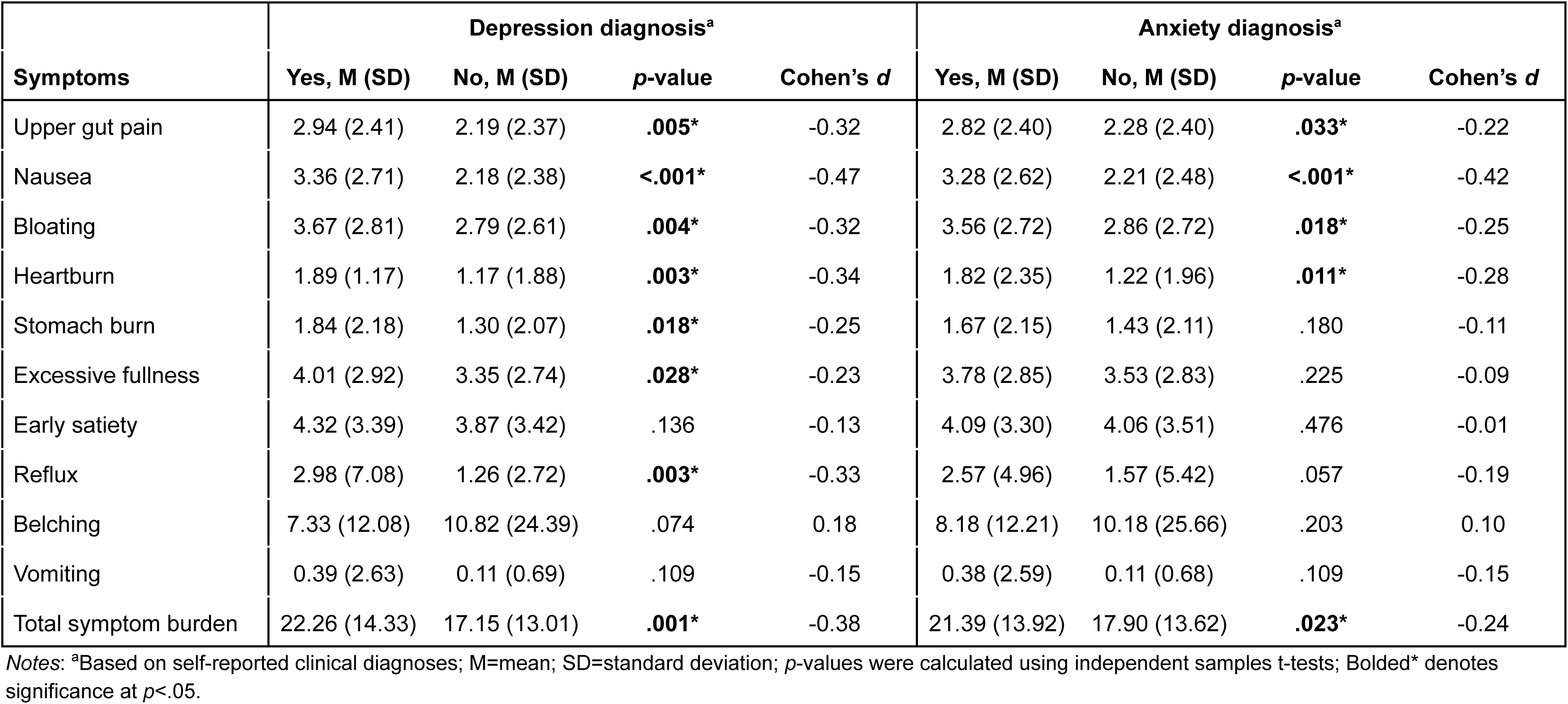
Independent samples t-tests comparing gastroduodenal symptoms between those patients with and without self-reported clinical diagnoses of depression and anxiety.

.30).

### Relationships across Rome groupings

A greater number of Rome diagnoses was associated with significantly poorer mental health outcomes. Specifically, patients meeting the criteria for both FD and CNVS exhibited significantly worse depression, anxiety, stress, and quality of life compared to those with FD alone (all *p*< .001) and significantly worse stress and quality of life than those with CNVS alone (all *p*< .031).

Furthermore, the relationships between symptoms and psychometrics varied across different diagnostic groups. Patients with CNVS or PDS alone showed no significant correlations between any psychometrics and symptoms (all *p*> .103), whilst patients with EPS alone demonstrated only a few significant correlations, specifically between anxiety and belching (*r*= .41, *p*= .032), and between stress and upper gut pain (*r*= .49, *p*= .010). Consistent with the full sample analysis, patients with overlapping CNVS and FD exhibited significant correlations between the majority of symptoms and psychometric measures.

### Metric-psychometric relationships

Pearson’s correlations showed no significant associations between the three BSGM metrics and depression, anxiety, stress, or quality of life (all *p*> .122), including when split by Rome diagnoses. Similarly, independent-sample t-tests demonstrated no significant differences in the BSGM metrics observed between those with and without a self-reported clinical diagnosis of anxiety or depression (all *p*> .074).

### BSGM Phenotypes

As shown in Figure 3, the majority of patients had normal spectral metrics with continuous and/or meal-induced symptom profiles.

**Figure 3.**
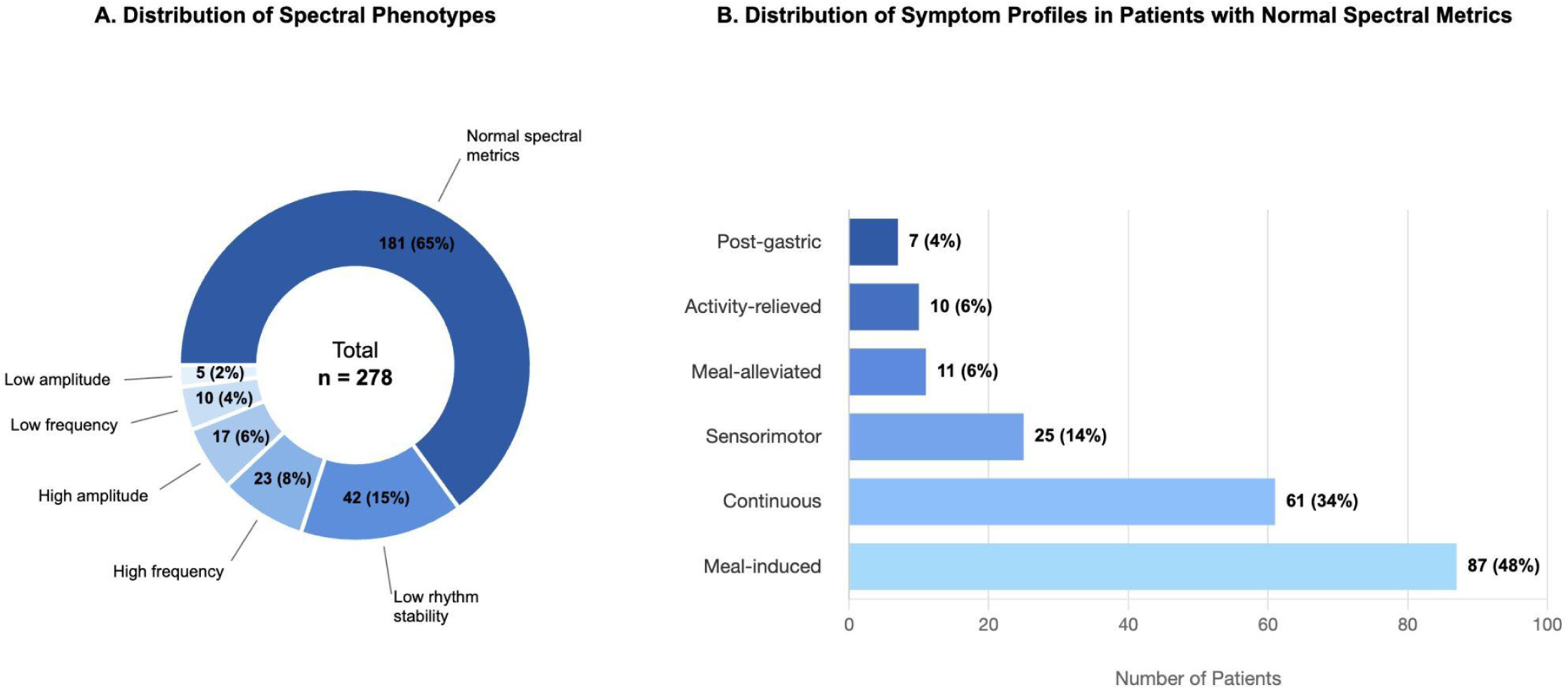
Distribution of (A) spectral phenotypes and (B) symptom profiles in patients with normal spectral metrics.

The regression analyses (see Table 2) revealed several significant associations between the BSGM phenotypes and the psychometrics. Having an abnormal rhythm stability (‘Dysrhythmic Phenotype’) significantly predicted higher depression (*p*= .044) and stress scores (*p*= .026), which was the only spectral phenotype associated with the psychometrics. Normal spectral metrics combined with continuous symptoms (‘Continuous Phenotype’) predicted higher depression (*p*= .003), anxiety (*p*= .045), and stress (*p*= .008), and lower quality of life (*p*< .001). Additionally, normal spectral metrics paired with sensorimotor symptoms (‘Sensorimotor Phenotype’) predicted higher anxiety (*p*= .029) and lower quality of life (*p*= .033). Lastly, normal spectral metrics combined with activity-relieved symptoms predicted higher stress (*p*= .043) and post-gastric symptoms predicted lower depression (*p*= .013).

**Table 2.**
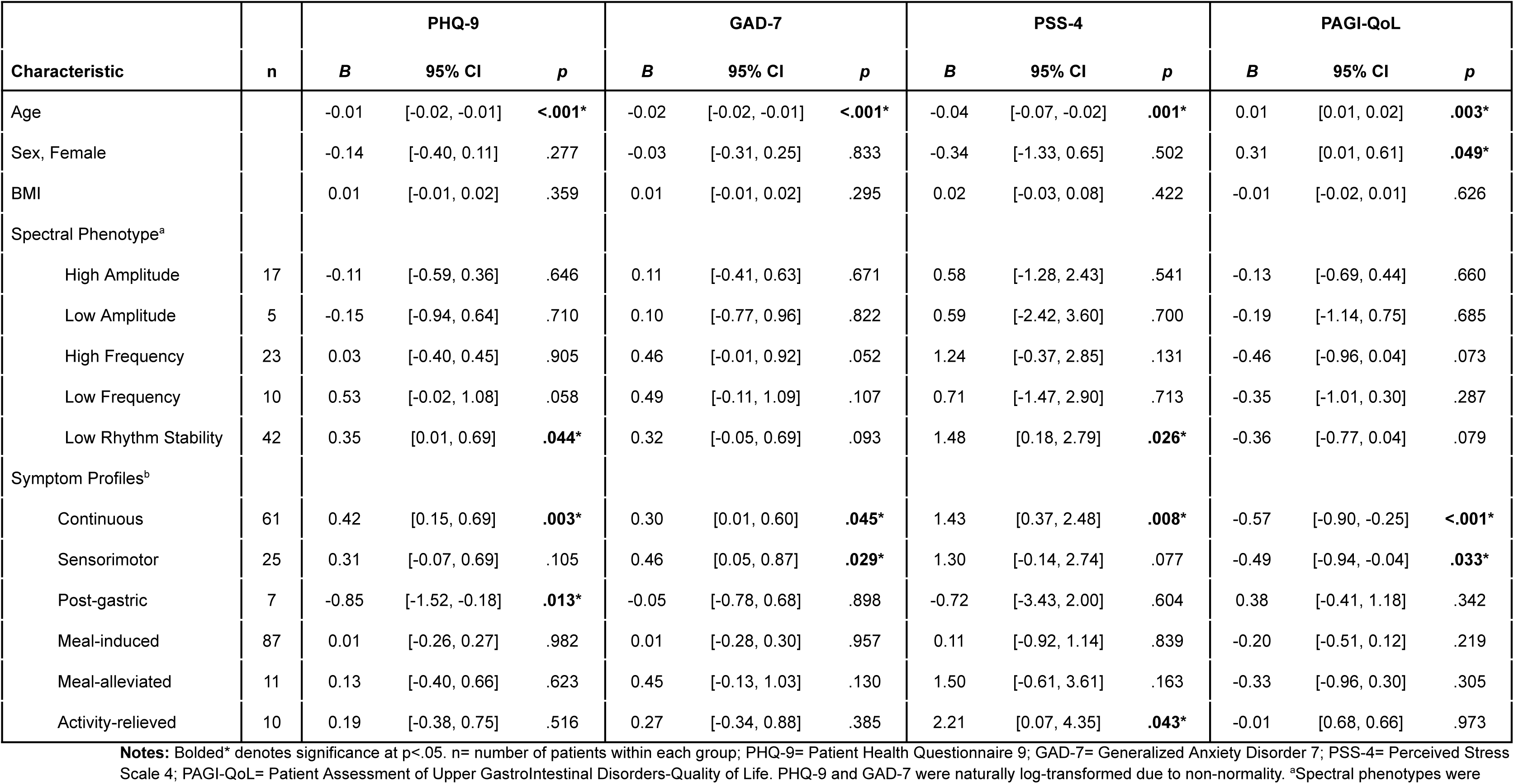
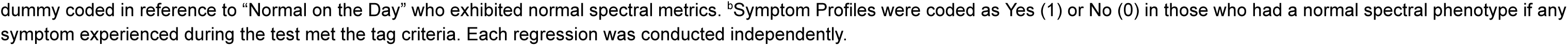
Regression statistics showing associations between the body surface gastric mapping phenotypes based on spectral metrics and symptom profiling, and depression (PHQ-9), anxiety (GAD-7), stress (PSS-4), and quality of life (PAGI-QoL).

| Characteristic | n | PHQ-9 |  |  | GAD-7 |  |  | PSS-4 |  |  | PAGI-QoL |  |  |
| --- | --- | --- | --- | --- | --- | --- | --- | --- | --- | --- | --- | --- | --- |
|  |  | *B* | 95% CI | *p* | *B* | 95% CI | *p* | *B* | 95% CI | *p* | *B* | 95% CI | *p* |
| Age |  | -0.01 | [-0.02, -0.01] | **<.001\*** | -0.02 | [-0.02, -0.01] | **<.001\*** | -0.04 | [-0.07, -0.02] | **.001\*** | 0.01 | [0.01, 0.02] | **.003\*** |
| Sex, Female |  | -0.14 | [-0.40, 0.11] | .277 | -0.03 | [-0.31, 0.25] | .833 | -0.34 | [-1.33, 0.65] | .502 | 0.31 | [0.01, 0.61] | **.049\*** |
| BMI |  | 0.01 | [-0.01, 0.02] | .359 | 0.01 | [-0.01, 0.02] | .295 | 0.02 | [-0.03, 0.08] | .422 | -0.01 | [-0.02, 0.01] | .626 |
| Spectral Phenotype<sup>a</sup> |  |  |  |  |  |  |  |  |  |  |  |  |  |
| High Amplitude | 17 | -0.11 | [-0.59, 0.36] | .646 | 0.11 | [-0.41, 0.63] | .671 | 0.58 | [-1.28, 2.43] | .541 | -0.13 | [-0.69, 0.44] | .660 |
| Low Amplitude | 5 | -0.15 | [-0.94, 0.64] | .710 | 0.10 | [-0.77, 0.96] | .822 | 0.59 | [-2.42, 3.60] | .700 | -0.19 | [-1.14, 0.75] | .685 |
| High Frequency | 23 | 0.03 | [-0.40, 0.45] | .905 | 0.46 | [-0.01, 0.92] | .052 | 1.24 | [-0.37, 2.85] | .131 | -0.46 | [-0.96, 0.04] | .073 |
| Low Frequency | 10 | 0.53 | [-0.02, 1.08] | .058 | 0.49 | [-0.11, 1.09] | .107 | 0.71 | [-1.47, 2.90] | .713 | -0.35 | [-1.01, 0.30] | .287 |
| Low Rhythm Stability | 42 | 0.35 | [0.01, 0.69] | **.044\*** | 0.32 | [-0.05, 0.69] | .093 | 1.48 | [0.18, 2.79] | **.026\*** | -0.36 | [-0.77, 0.04] | .079 |
| Symptom Profiles<sup>b</sup> |  |  |  |  |  |  |  |  |  |  |  |  |  |
| Continuous | 61 | 0.42 | [0.15, 0.69] | **.003\*** | 0.30 | [0.01, 0.60] | **.045\*** | 1.43 | [0.37, 2.48] | **.008\*** | -0.57 | [-0.90, -0.25] | **<.001\*** |
| Sensorimotor | 25 | 0.31 | [-0.07, 0.69] | .105 | 0.46 | [0.05, 0.87] | **.029\*** | 1.30 | [-0.14, 2.74] | .077 | -0.49 | [-0.94, -0.04] | **.033\*** |
| Post-gastric | 7 | -0.85 | [-1.52, -0.18] | **.013\*** | -0.05 | [-0.78, 0.68] | .898 | -0.72 | [-3.43, 2.00] | .604 | 0.38 | [-0.41, 1.18] | .342 |
| Meal-induced | 87 | 0.01 | [-0.26, 0.27] | .982 | 0.01 | [-0.28, 0.30] | .957 | 0.11 | [-0.92, 1.14] | .839 | -0.20 | [-0.51, 0.12] | .219 |
| Meal-alleviated | 11 | 0.13 | [-0.40, 0.66] | .623 | 0.45 | [-0.13, 1.03] | .130 | 1.50 | [-0.61, 3.61] | .163 | -0.33 | [-0.96, 0.30] | .305 |
| Activity-relieved | 10 | 0.19 | [-0.38, 0.75] | .516 | 0.27 | [-0.34, 0.88] | .385 | 2.21 | [0.07, 4.35] | **.043\*** | -0.01 | [0.68, 0.66] | .973 |

**Notes:** Bolded\* denotes significance at $p < .05$ . n= number of patients within each group; PHQ-9= Patient Health Questionnaire 9; GAD-7= Generalized Anxiety Disorder 7; PSS-4= Perceived Stress Scale 4; PAGI-QoL= Patient Assessment of Upper Gastrointestinal Disorders-Quality of Life. PHQ-9 and GAD-7 were naturally log-transformed due to non-normality. <sup>a</sup>Spectral phenotypes were
dummy coded in reference to “Normal on the Day” who exhibited normal spectral metrics. <sup>b</sup>Symptom Profiles were coded as Yes (1) or No (0) in those who had a normal spectral phenotype if any symptom experienced during the test met the tag criteria. Each regression was conducted independently.

## Discussion

This international BSGM consortium study examined the associations between psychological factors and emerging BSGM test phenotypes in patients with FD and CNVS. As anticipated, our patient cohort exhibited considerably elevated levels of depression, anxiety, and stress compared to those observed in the general population^38–40^; with levels of clinical diagnoses over 40% higher than the general population^41^. These psychological variables were significantly associated with symptom severity, particularly in patients with overlapping FD and CNVS. This aligns with literature indicating that patients with overlapping DGBI diagnoses often experience worse mental health and gastrointestinal symptoms^42,43^. However, the observed psychological associations varied systematically according to BSGM-defined phenotypes, rather than reflecting a generic psychological overlay.

The regression analyses performed here reveal distinct psychometric profiles across the BSGM disease phenotypes, supporting the utility of this emerging classification system for identifying clinically meaningful patient subgroups. Specifically, having an abnormal rhythm stability, indicated by a low GA-RI (‘Dysrhythmic Phenotype’), predicted higher levels of depression, potentially reflecting the debilitating impact of more severe gastric dysregulation on mental health. A similar observation was recently observed in a separate adolescent population with abnormal rhythms, who also showed more severe symptoms and reduced quality of life^24^. While the directionality of this relationship now warrants further investigation, it highlights the importance of measuring and addressing mental health, even in patients with significant gastric dysfunction. Within the current BSGM classification hierarchy, dysrhythmia warrants primary pathophysiological investigation and management, with psychological support serving as a complementary, though important consideration^27^.

In contrast, abnormal Principal Gastric Frequency and BMI-Adjusted Amplitude did not exhibit any significant relationships with the measured psychological variables. This suggests that these abnormalities may be less indicative of gut-brain dysfunction and more likely to reflect underlying gastric motility disturbances, gastric outlet resistance, vagal nerve dysfunction, or gastric hypomotility, as supported by previous studies^14,17,18,44^.

Conversely, multiple significant associations emerged between the psychometrics and symptom patterns in specific patient subgroups with normal spectral metrics. This aligns with prior research demonstrating that individuals with normal spectral metrics on BSGM displayed higher levels of depression and anxiety than those with identifiable gastric abnormalities on spectral analysis^14,45^. Notably, patients with normal spectral metrics and continuous symptom patterns (‘Continuous Phenotype’) predicted significantly higher depression, anxiety, and stress, and lower quality of life; the only group to have relationships with all four psychological variables measured. The lack of association between symptoms and gastric activity in this subgroup, coupled with high psychological comorbidities, suggests gut-brain dysregulation as a contributing symptom driver, potentially in tandem with other mediators such as chronic inflammation^46^. This particular BSGM pattern could serve as a proxy for a clinical diagnosis of the EPS subtype of FD^28^, suggesting that BSGM testing could offer a more robust detection tool than symptom-based criteria alone.

Furthermore, normal spectral metrics combined with sensorimotor symptoms (‘Sensorimotor Phenotype’) were associated with higher anxiety and lower quality of life. We hypothesize that pathophysiology in this subgroup may be linked to visceral hypersensitivity, where normal digestive sensations are perceived as painful or symptomatic, potentially leading to food-related anxiety or altered interoception^47,48^. This finding aligns with hypotheses generated from previous research in patients with sensorimotor symptoms^19,23,27,49^.

Consequently, we hypothesize that this group may benefit from interventions targeting visceral hypersensitivity and interoception, including neuromodulators, vagal stimulation, or psychological therapies aimed at modifying symptom perception^50^. This particular BSGM pattern could also serve as a proxy for a clinical diagnosis of the PDS subtype of FD^28^; offering the additional advantage of ruling out neuromuscular dysfunction via a normal spectral phenotype.

While other symptom profiles, such as activity-relieved and post-gastric, showed associations with depression and stress, these phenotypes remain uncommon in our database, and as such, these findings require cautious interpretation. Further work is required to define the clinical utility of these symptom patterns. Additionally, the most prevalent symptom tag, meal-induced, comprising 31% of the sample, exhibited no significant associations with any of the psychometric measures. This suggests that this particular symptom pattern may not be associated with neither gut-brain dysregulation nor gastric myoelectrical dysfunction, potentially implicating other known gastroduodenal pathophysiologies not measured in this study, such as immune activation^51,52^. Further investigation into this subgroup is warranted to elucidate the mechanisms underlying their meal-induced symptomatology.

Overall, these findings support the value of integrating BSGM metrics, symptom profiles, and psychological assessments to enhance the precision of patient phenotyping, in gastroduodenal disorders. We anticipate that this approach could enable more tailored, mechanism-informed management strategies. Specifically, we hypothesize that patients with normal spectral metrics and continuous or sensorimotor symptoms may receive greater benefit from multidisciplinary approaches incorporating psychological interventions, such as CBT, hypnotherapy, or biofeedback, which have demonstrated efficacy in addressing both gastrointestinal symptoms and mental health^10,11,53^. Conversely, patients exhibiting spectral abnormalities may be more responsive to targeted medical approaches, including pharmacological interventions and surgical procedures^21^. Studies have indicated that this personalized approach, guided by BSGM profiling, can lead to improved healthcare utilization and cost reductions^20,54^. In addition, guiding patients towards appropriate psychological interventions is crucial given the limited access to these therapies^55^. Additional studies are ongoing to further assess these relationships.

The current findings further support the recent integration of the Alimetry Gut-Brain Wellbeing (AGBW) Survey^56^ into the standard Gastric Alimetry BSGM test protocol, in order to offer a more holistic assessment of patients with gastroduodenal disorders. This validated tool evaluates depression, anxiety, and stress in patients with chronic gastroduodenal symptoms. Future research is expected to leverage the AGBW Survey to further evaluate the associations between its scores and the BSGM metrics, symptom patterns, and phenotypes.

This study has several limitations. Firstly, the observational design precludes causal inference between BSGM phenotypes and mental health outcomes. Longitudinal and interventional trials would be needed to confirm the proposed mechanisms and delineate the temporal relationship between symptoms and psychological comorbidity, though such studies are resource-intensive and logistically challenging. Secondly, the Western countries sampled, although international, may limit the global generalizability of our findings. Lastly, despite the relatively large sample size, certain phenotypes and symptom profiles were uncommon, potentially affecting the accuracy of the psychological profiling in these subgroups. Further studies with larger cohorts and targeted recruitment would be needed to ensure adequate representation of such patient subgroups. Alternatively, if these subgroups continue to offer marginal clinical benefit, they may be excluded from future classification systems to enhance clarity and practicality.

In summary, this study shows substantial psychological comorbidity in chronic gastroduodenal disorders, which is not uniform but varies strongly by BSGM phenotypes. Psychological burden was highest in patients with normal gastric rhythms and continuous or sensorimotor symptoms, suggesting these groups are most likely to benefit from psychological interventions. These findings support the role of multimodal BSGM testing protocols in identifying specific phenotypes to guide more targeted, integrative care.

## Data Availability

Deidentified data will be made available upon reasonable request to the corresponding author.

## Disclosures

GOG and AG hold intellectual property and grants in gastric electrophysiology and are Directors of the University of Auckland spin-out company Alimetry Ltd. ML, GS, BW, CD, DF, CNA, and SC are members of Alimetry Ltd. The remaining authors have no relevant conflicts to declare.

## Grant Support

This study is funded by a New Zealand Health Research Council Programme Grant 3715588.

## Data transparency statement

Deidentified data will be made available upon reasonable request to the corresponding author.

## Notes

### Author Declarations

Ethical approvals were granted by the Auckland Health Research Ethics Committee (AHREC; AH1130 & AH27068), the University of Calgary Conjoint Health Research Ethics Board (REB19-1925), and the Human Research Ethics Committee at Western Sydney (H15157). All participants provided written informed consent.

